# Novel Ripple Frequency–Guided Substrate Targeting for Persistent Atrial Fibrillation: 12-Month Outcomes

**DOI:** 10.1101/2025.09.25.25336693

**Authors:** Daniel P Melby, Venkatakrishna N Tholakanahalli, Refael Itah, Robert G Hauser, Sean Mazer, Edwin T Zishiri, Jay D Sengupta, Matthew D Olson, Emily Halvorson, Ali Bahbah, Joseph J Decker, Charles C Gornick, David G Benditt

## Abstract

**Background:** Outcomes after pulmonary vein isolation (PVI) for persistent atrial fibrillation (AF) remain limited, motivating patient-specific strategies to identify non-PV sources. Ripple Frequency is an automated mapping algorithm designed to localize AF sources for targeted ablation by highlighting regions with high frequency directional changes in electrogram dV/dT.

**Objective:** To evaluate 12-month freedom from AF and any atrial arrhythmia following non-PV Ripple Frequency–guided ablation for persistent AF.

**Methods:** We analyzed 72 patients undergoing first-time ablation for persistent AF. After PVI, ablation was directed at non-PV atrial regions in the top quartile of each patient’s Ripple Frequency maps. The primary endpoint was first AF recurrence >30 s after a 90-day blanking period on or off antiarrhythmic drugs (AADs); secondary endpoints included atrial tachycardia/flutter (AT/AFL), any atrial arrhythmia, and safety. Freedom from AF at 12 months was compared with that observed in the PVI arm of the STAR-AF II trial for additional context.

**Results:** Ripple Frequency targets were ablated in 66/72 patients (92%), yielding acute AF termination in 64/72 patients (88.9%). Single procedure 12-month freedom from AF was 95.8% (95% CI 87.6-98.6) on/off AAD, and freedom from any atrial arrhythmia was 72.2% (95% CI 60.3-81.1). After 1.2 procedures, these rose to 97.2% and 90.3%. Compared with the STAR-AF II PVI arm (60%), 12-month AF freedom was 35.8% (95% CI 22.3-40.0%) higher.

**Conclusion:** In this single-center cohort, non-PV ablation guided by Ripple Frequency was associated with a high 12-month AF and any arrhythmia freedom with no major complications.

**Central Illustration:** 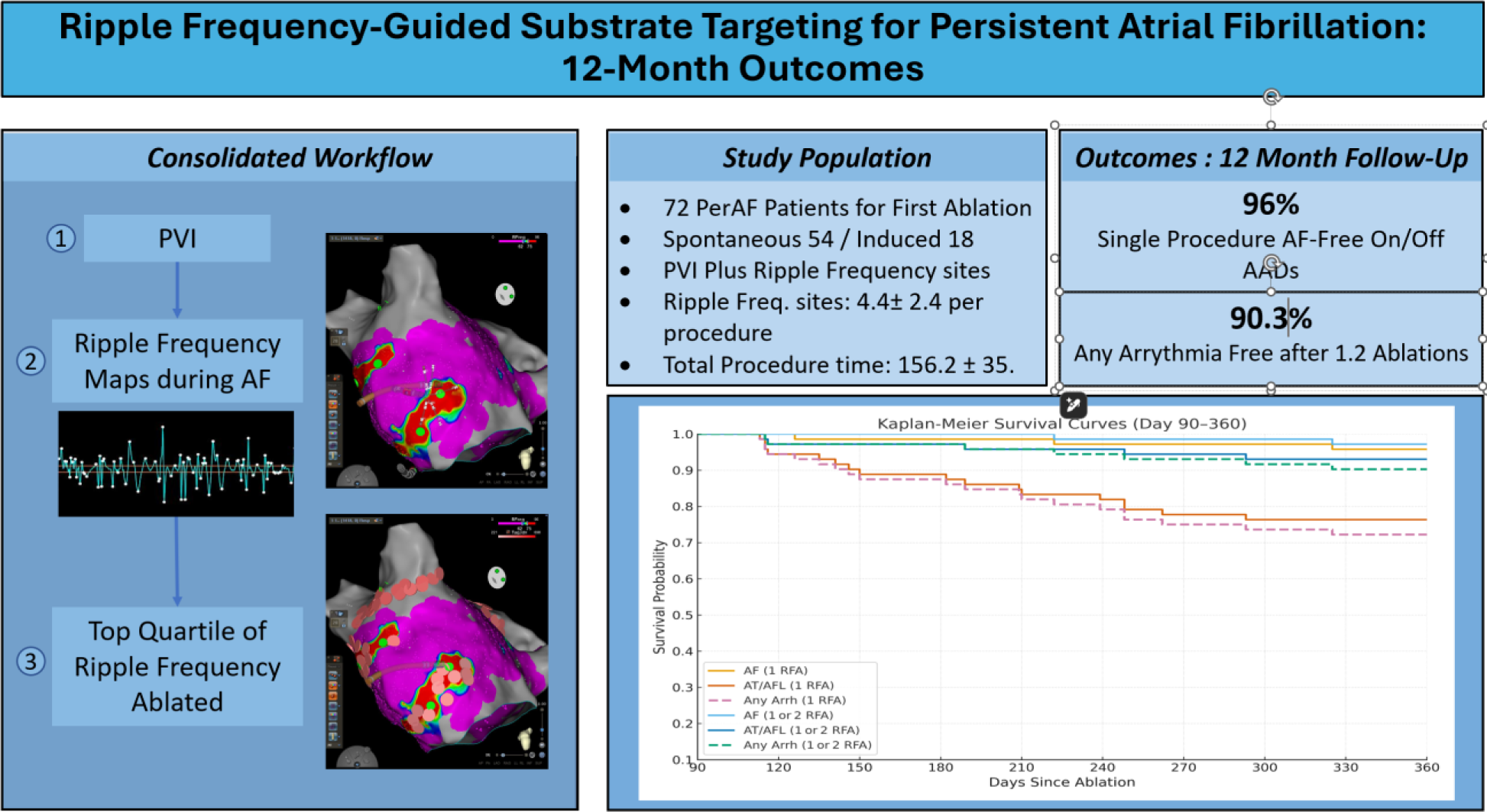

## 1. Introduction

Atrial fibrillation (AF) is the most common sustained cardiac arrhythmia encountered in clinical practice and an important contributor to morbidity and health care utilization. (1) Catheter ablation, most commonly pulmonary vein isolation (PVI), is an established therapy for symptomatic patients who do not tolerate or respond to antiarrhythmic drugs. In persistent AF, however, single-procedure outcomes after PVI alone remain modest (≈60% arrhythmia-free survival at 1 year), with recurrences attributed to atrial substrate and non–pulmonary-vein (PV) triggers. Limited evidence defines optimal strategies for identifying and targeting such non-PV sources (2,3), sustaining interest in mapping approaches that can support patient-specific ablation beyond PVI.

Ripple Frequency is an automated mapping tool that integrates multiple electrograms within a regional window to detect areas where high-frequency changes in dV/dT occur. These electrogram findings may reflect wavefront crowding, re-entry, or unstable conduction pathways consistent with potential AF source locations. Building on prior work (4,5), we evaluated Ripple Frequency–guided ablation performed in real time after wide-area PVI within a single-center cohort. The prespecified objective was to assess 12-month freedom from AF, with secondary endpoints including other atrial arrhythmias and safety; outcomes are also contextualized against historical PVI-only data.

## 2. Methods

### 2.1 Study Design and Population

We performed a single-center retrospective cohort study. Ripple Frequency mapping was applied in real time during each procedure between September 2023 and January 2024, but all outcome extraction and statistical analyses were conducted retrospectively. Consecutive patients undergoing first-time ablation for persistent atrial fibrillation (continuous AF > 7 days duration) who met the following criteria were included in the study: age of 18 years or older, documented persistent AF on a multiday ambulatory ECG, an ambulatory rhythm monitor, or an implanted cardiac device, class I or II guideline eligibility for catheter ablation (1), and consent for participation in research. Exclusion criteria included prior ablation (surgical or catheter) for AF, severe LV dysfunction (EF <25%), structural heart disease precluding ablation, left atrial thrombus, contraindication to anticoagulation, and pregnancy. The Ripple Frequency software obtained FDA 510(k) approval (K221112). The Allina Health Institutional Review Board, Minneapolis, Minnesota, approved this study (IRB # 2083021).

The primary endpoint was the first documented recurrence of AF >30 seconds in duration on/off AADs, detected on either a 12-lead ECG, ambulatory monitor, or continuous cardiac implantable electronic device (CIED) interrogation after a 90-day blanking period. (1) Secondary endpoints were (i) first recurrence of atrial tachycardia or atrial flutter (AT/AFL) >30 seconds in duration, and (ii) occurrence of any atrial arrhythmia (AF, AT, or AFL). Early recurrences during days 0–90 (blanking period) were managed clinically but were not counted as endpoint events. Participants were followed until first documented arrhythmia (>30 s) after the 90-day blanking period; those without recurrence were administratively censored at day 360.

### 2.2 Real-Time Identification of Ripple Frequency Sites

Ripple Frequency is a novel Carto 3 algorithm that measures, during a 2.5 second signal acquisition window, the number of directional changes in dV/dT per second. This method detects regions of highly fractionated electrograms that may correspond to AF sources. (Figure 1) To minimize identification of very small anatomic locations or isolated single electrograms the algorithm uses wide regional averaging and voltage gate techniques.

**Figure 1:**
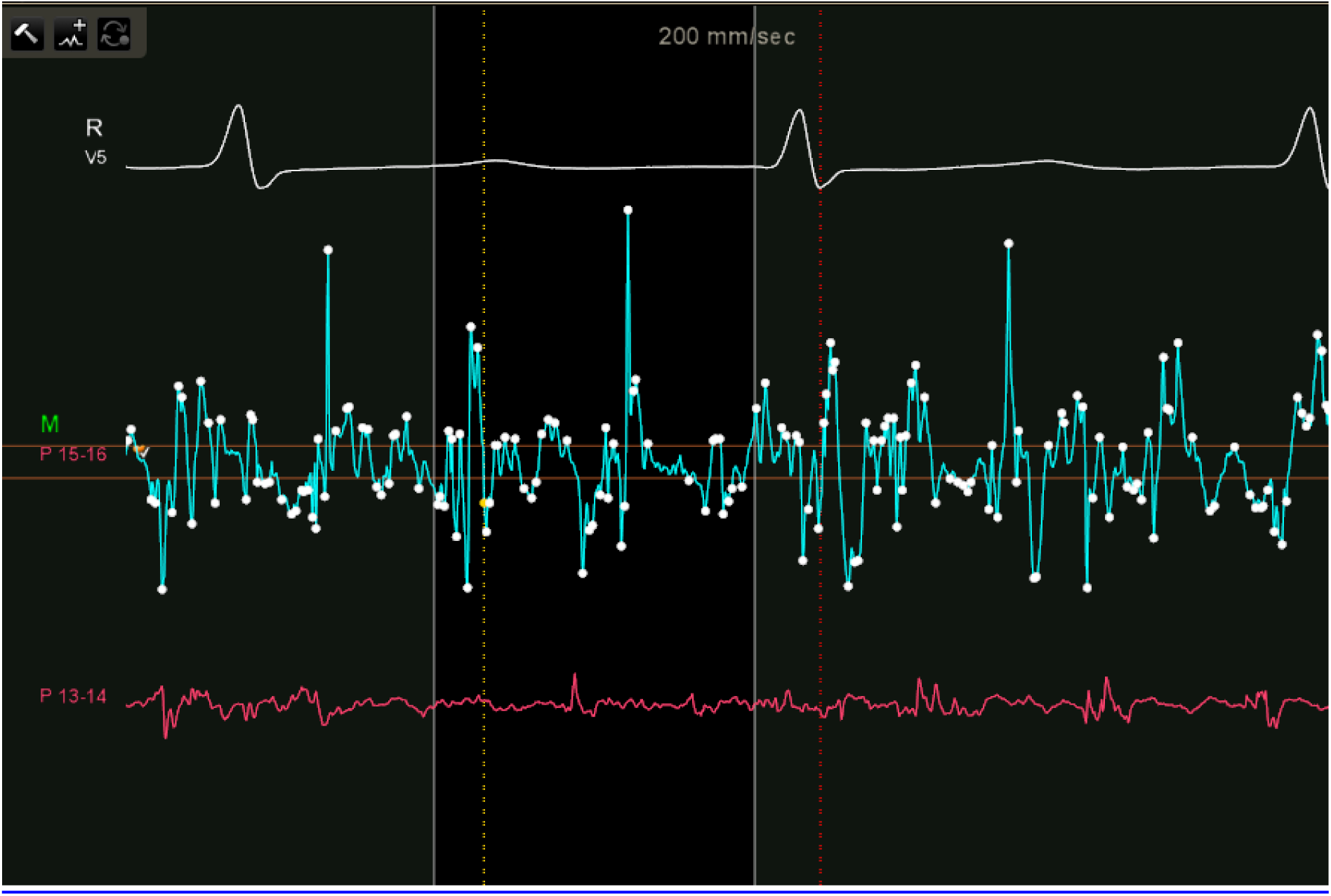
Example of Ripple Frequency algorithm. markers on an electrogram obtained during AF. The white markers represent a Ripple peak that was counted by the algorithm. The orange lines delineate the pre-set voltage thresholds of ± 0.03 mV from baseline 0 mV.

This study implemented real-time intra-procedural analysis of Ripple Frequency regions to guide non-PV ablation sites after completion of PVI. The upper quartile of Ripple Frequency, representing the highest 25% of Ripple Frequencies recorded in each atrium, was identified automatically via the software as potential ablation targets (Figure 2). This quartile threshold was dynamically adjusted based on the full dataset of Ripple Frequency for each patient to allow patient-specific AF source targeting. Our prior study (5) demonstrated that the upper quartile of Ripple Frequency had the highest sensitivity and specificity for detecting regions corresponding with visual Ripple map analysis.

**Figure 2:**
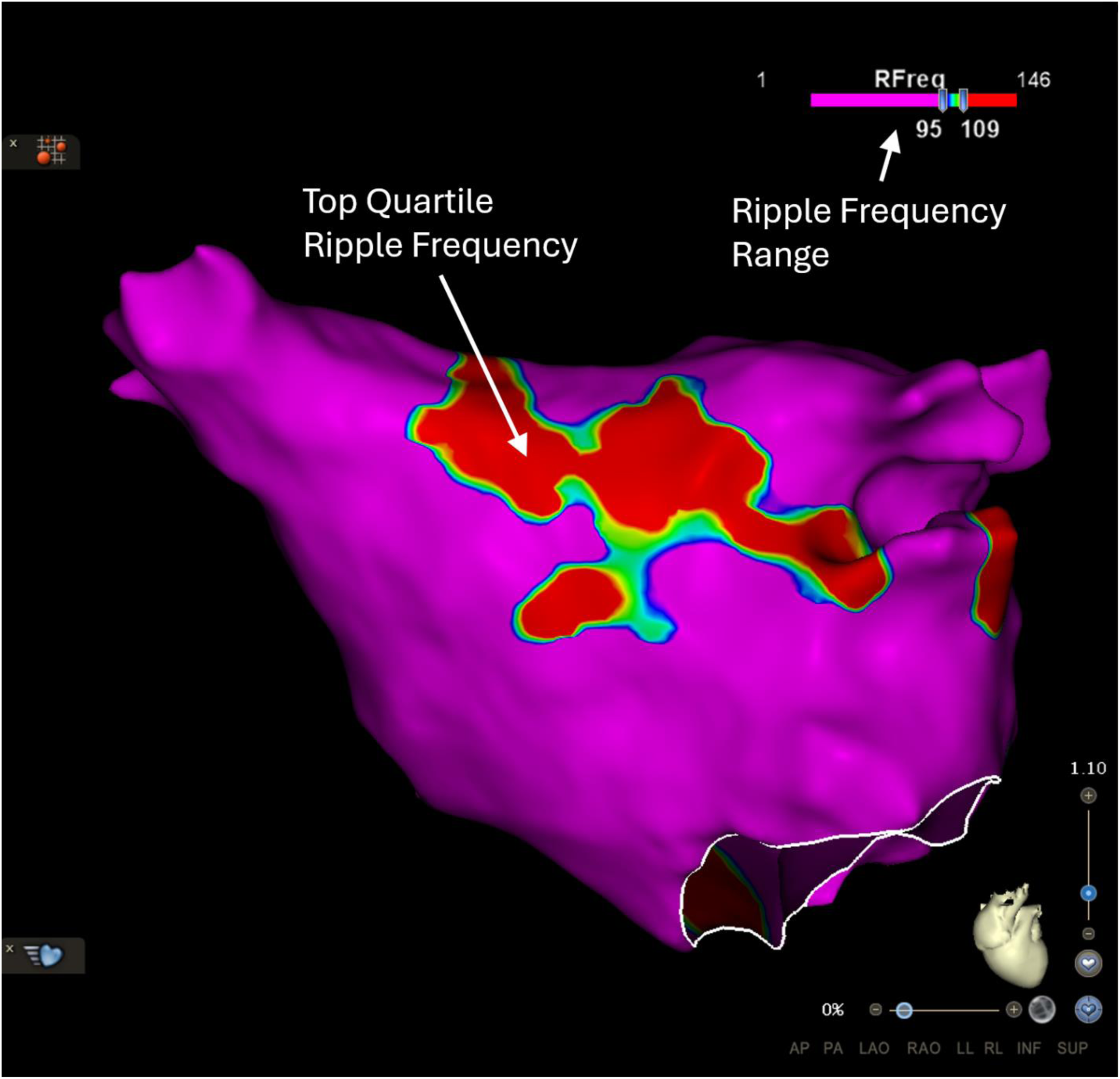
Example of Ripple Frequency map. obtained during sustained AF. Color shading represents the Ripple Frequency. Color bar (upper right) shows absolute Ripple Frequency values in s⁻¹; ticks correspond to 65%, and 75% of the maximum frequency recorded in this map (1–146 s⁻¹ in this case). The red color represents Ripple Frequency ≥75% of the maximum (in this case, 109 s⁻¹). The purple color represents Ripple Frequency <65% of the maximum (in this case, 95 s⁻¹). Analysis of this map demonstrated the upper quartile of Ripple Frequency concentrated at LA roof and extending to LSPV antrum, LA septum/anterior, and superior lateral ridge.

### 2.3 Electrophysiological Study and Mapping

The electroanatomic study and Ripple Frequency map were performed in all patients during AF. Patients who presented in normal sinus rhythm (NSR) at baseline underwent AF induction via atrial pacing from the coronary sinus. Left atrial (LA) and right atrial (RA) maps were constructed using the Carto 3 Confidense algorithm with a Pentaray or Octaray catheter (J&J MedTech, 2-6-2 mm or 2-2-2-2-2 mm electrode spacing). Intracardiac bipolar electrograms were acquired with the Carto 3 recording system. Signals were band-pass filtered 16–500 Hz (high-pass 16 Hz, low-pass 500 Hz). Electroanatomic point density was set to 1 mm to enable dense regional point acquisition. Electrode stability was maintained using a novel Confidense algorithm filter (J&J MedTech). Points with electrode motion >3 mm (SD) during the 2.5-s acquisition window were automatically excluded to diminish movement artifact.

Our prespecified workflow required: (1) automatic Ripple Frequency target generation without manual editing, (2) ≥1,500 left atrial and ≥1,000 right atrial mapping points, and (3) real-time lesion delivery following the protocolized sequence outlined below. Mapping logs were automatically exported and archived for independent audit. Antiarrhythmic medications were held for five drug half-lives (except one week for amiodarone) before the ablation procedure. All procedures were performed under general anesthesia with continuous hemodynamic and esophageal temperature monitoring. A low fluoroscopy workflow was performed during these procedures.

### 2.4 Ablation Protocol

PVI was completed in all patients with an open-irrigated, contact-force–sensing catheter (THERMOCOOL SMARTTOUCH SF or QDOT Micro, J&J MedTech). After PV entrance block was confirmed, if AF did not terminate with PVI, the left atrium (LA) was remapped as many as three times. Atrial regions within the top quartile of Ripple Frequency were ablated sequentially, beginning with confluent areas larger than 1 cm² and progressing to smaller foci until atrial fibrillation (AF) terminated. Ablation lesions were delivered as contiguous clusters and expanded until local cycle-length prolongation, electrogram attenuation, or arrhythmia termination was observed. If AF persisted, the right atrium (RA) was mapped, and Ripple Frequency targets were ablated in the same manner.

Radio-frequency energy was applied at 35–50 W with ≥ 5 g contact force; 40 W was customary for linear lesions (e.g., LA roof, mitral isthmus). Energy applications along the posterior LA adjacent to the esophagus were shortened to minimize thermal injury. The procedural endpoint was AF termination; if AF or multifocal atrial tachycardia (MAT) remained, electrical cardioversion was performed. MAT was defined as varying biatrial activation patterns, biatrial absence of fibrillatory electrograms, and biatrial cycle lengths exceeding 200 ms. Additional linear lesions were delivered only for sustained macro-reentrant tachycardia from AF termination or induction with atrial pacing. Before procedure completion, atrial burst pacing (short duration, 3-5 seconds), to a cycle length of 230 ms was performed to assess for readily inducible AF or AT/AFL. If either arrhythmia was initiated, ablation was performed until arrhythmia termination (or cardioversion if needed), and repeat induction failed to reinduce arrhythmia.

### 2.5 Post-Procedural Monitoring

Rhythm monitoring was conducted at the time of clinical follow-up at the discretion of the treating electrophysiologist. Patient follow-up to assess AF ablation success was determined by standard techniques, which included electrocardiograms, outpatient rhythm monitors, CIED and/or symptom-directed event monitoring. Oral anticoagulation was continued post-procedure per guideline recommendations, and antiarrhythmic drug (AAD) therapy was maintained or discontinued at the discretion of the attending physician. Two cardiologists adjudicated all qualifying rhythm recordings; discrepancies were resolved by consensus.

### 2.6 Safety Endpoints and Surveillance

Procedural and post-procedural 30-day adverse events were prospectively captured from catheter-laboratory logs and electronic health record reviews. Major procedure-related complications were prospectively predefined as per HRS/ECAS consensus (6): (i) cardiac tamponade requiring pericardiocentesis or surgery, (ii) stroke or transient ischemic attack, (iii) clinically significant esophageal injury (endoscopically proven ulcer or fistula), (iv) phrenic nerve palsy persisting for more than 24 hours, (v) vascular complication necessitating surgical or transfusion therapy, and (vi) procedure-related death within 30 days.

### 2.7 Statistical Analysis

All continuous variables were reported as mean ± standard deviation, and categorical variables as counts (%). We anticipated a 12-month single-procedure AF-free rate of 60% after PVI alone (2) and hypothesized a 25-percentage-point improvement with Ripple Frequency targeting. With α = 0.05 and 80% power, an intentionally conservative sample size of 66 patients was chosen; we enrolled 72 to offset a 10% attrition rate. Following the 90-day blanking period, participants were censored at the time of first documented arrhythmia (>30 seconds). Two-sided 95% confidence intervals (CIs) were calculated with Greenwood’s formula. Time-to-event curves began on day 91 (end of blanking). Participants without documented arrhythmia recurrence were censored on day 360.

#### Benchmark comparison

To contextualize single-arm outcomes, we compared our 12-month freedom-from-AF rate with the PVI arm of the STAR-AF II trial, the most extensive contemporary single-procedure series in persistent AF. (2) The Kaplan–Meier curve from STAR-AF II was digitized (Engauge Digitizer v12); patient-level survival data were reconstructed with the Guyot algorithm. (7) Absolute risk difference (RD) with 95 % confidence interval (CI) was calculated using the Newcombe method (8). This comparison was exploratory and subject to era and population-based differences.

## 3. Results

### 3.1 Participant Flow

During the enrollment period (September 2023 to January 2024), 77 consecutive patients with persistent AF were screened and underwent catheter ablation at our center. Five patients (6.5 %) were lost to follow-up and excluded from further analysis, yielding a final study cohort of 72 participants. All 72 patients completed at least one scheduled rhythm-surveillance evaluation following the 90-day blanking period, and none were censored before the 12-month endpoint due to withdrawal of consent. Accordingly, the entire cohort of 72 patients was included in the Kaplan–Meier analysis.

### 3.2 Baseline Demographics

Baseline demographic characteristics and echocardiographic findings of the study cohort are summarized in Table 1. Most patients were male (80.6%) with a mean age of 65.5±10.4 years. The mean body mass index (BMI) was 32.1 ± 6.8 kg/m². The mean CHA₂ DS₂ -VASc score was 2.3 ± 1.5. Echocardiography demonstrated a mean left atrial (LA) diameter of 45.8 ± 7.4 mm.

**Table 1.**
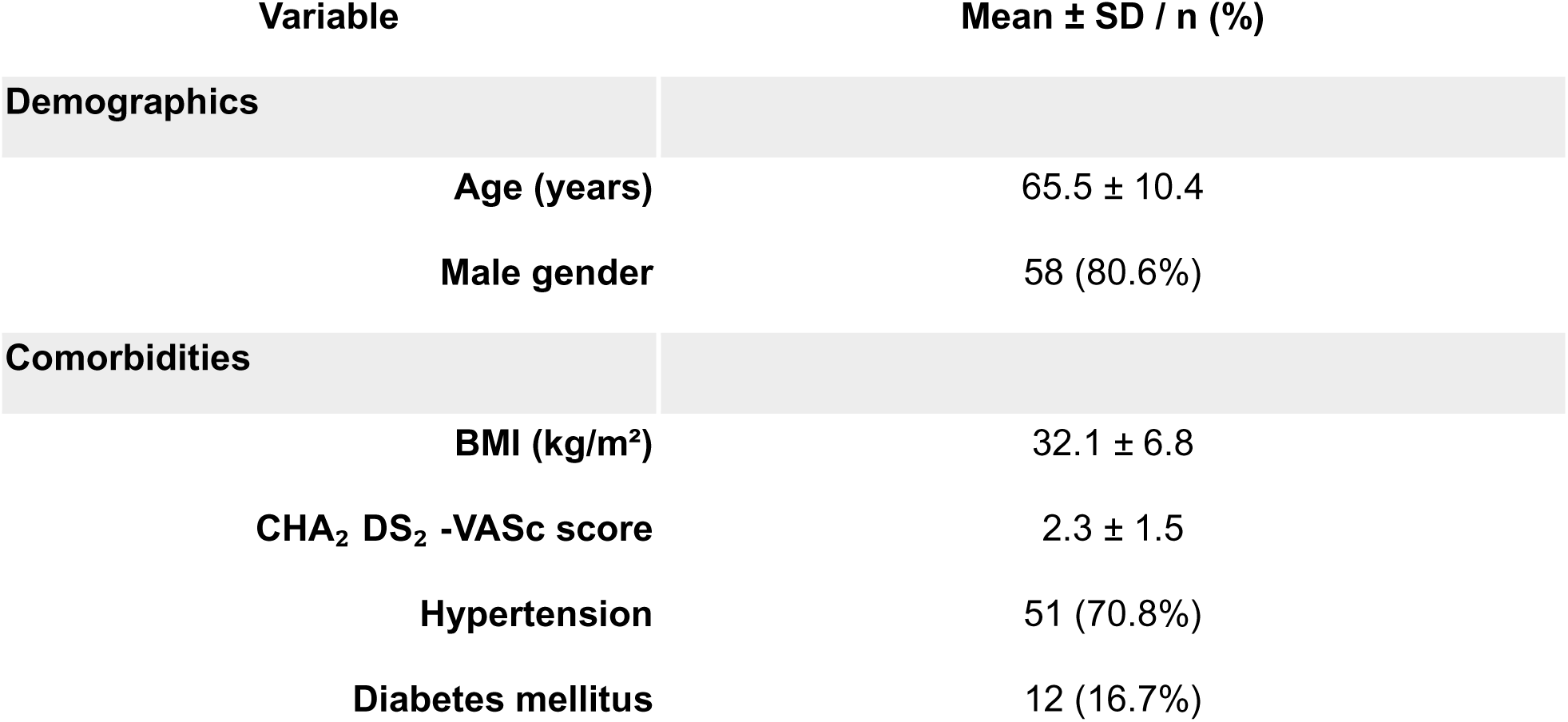
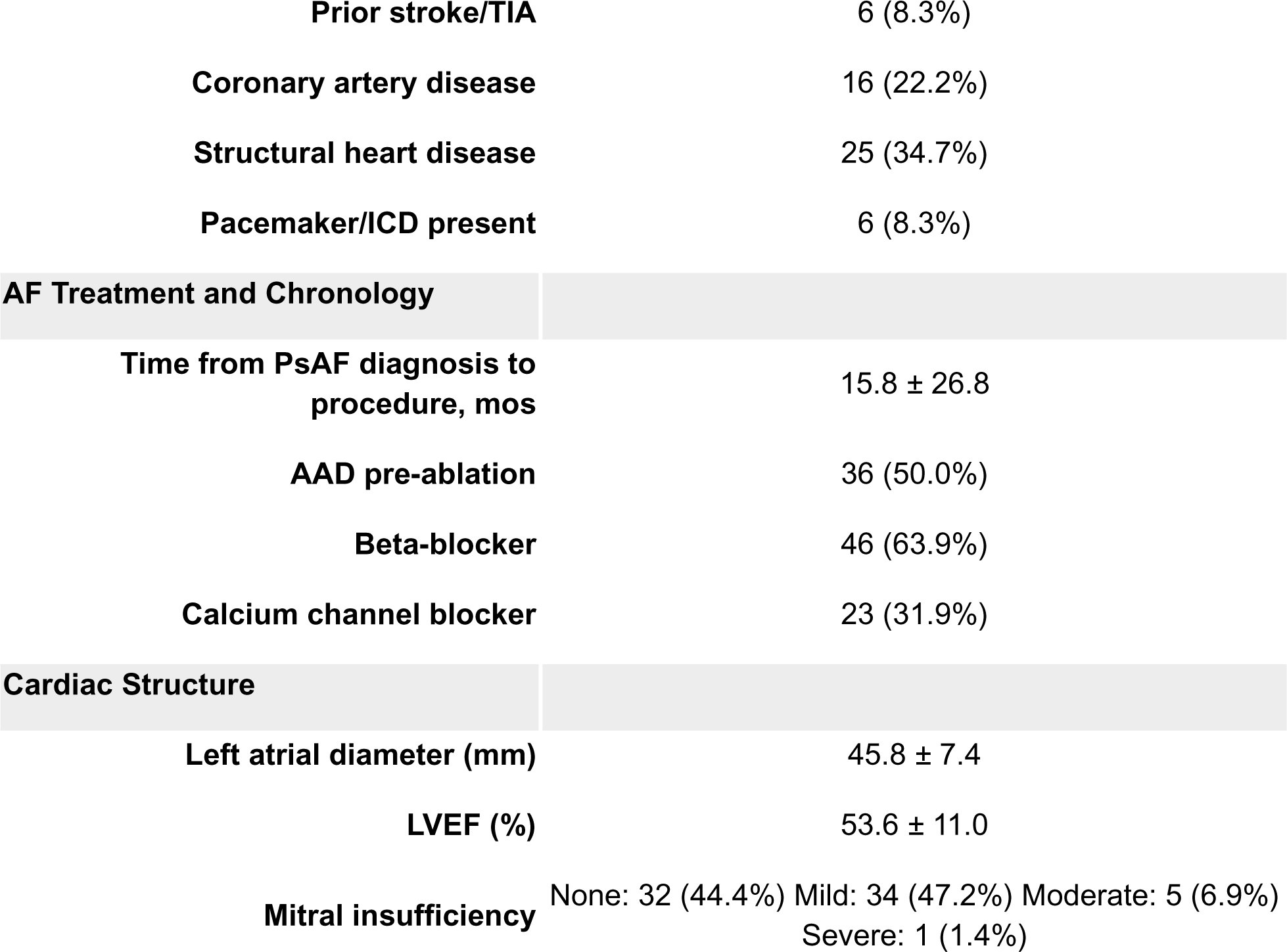
Baseline Characteristics of the Study Population (n = 72)

| Variable | Mean ± SD / n (%) |
| --- | --- |
| **Demographics** |  |
| Age (years) | 65.5 ± 10.4 |
| Male gender | 58 (80.6%) |
| **Comorbidities** |  |
| BMI (kg/m<sup>2</sup>) | 32.1 ± 6.8 |
| CHA<sub>2</sub> DS<sub>2</sub> -VASc score | 2.3 ± 1.5 |
| Hypertension | 51 (70.8%) |
| Diabetes mellitus | 12 (16.7%) |
| Prior stroke/TIA | 6 (8.3%) |
| Coronary artery disease | 16 (22.2%) |
| Structural heart disease | 25 (34.7%) |
| Pacemaker/ICD present | 6 (8.3%) |
| **AF Treatment and Chronology** |  |
| Time from PsAF diagnosis to procedure, mos | 15.8 ± 26.8 |
| AAD pre-ablation | 36 (50.0%) |
| Beta-blocker | 46 (63.9%) |
| Calcium channel blocker | 23 (31.9%) |
| **Cardiac Structure** |  |
| Left atrial diameter (mm) | 45.8 ± 7.4 |
| LVEF (%) | 53.6 ± 11.0 |
| Mitral insufficiency | None: 32 (44.4%) Mild: 34 (47.2%) Moderate: 5 (6.9%)<br>Severe: 1 (1.4%) |

### 3.3 Procedure Results

All patients underwent PVI. Electroanatomic mapping measured an enlarged mean LA volume of 160.0±39.4 mL. The mean baseline left atrial pressure was elevated (13.4 ± 4.4 mmHg). AF was pace-induced in 18 patients (25.0%), while the remainder presented in spontaneous AF. AF termination with PVI alone was observed in 6 patients (8.3%); the remaining 66 (91.7%) underwent additional Ripple Frequency specific ablation beyond the pulmonary veins. In total, AF termination (to NSR/Atrial tachycardia (AT)/ Atrial flutter (AFL) / Multifocal atrial tachycardia (MAT)) was observed in 64 patients (88.9%). Termination directly to NSR was observed in 11 patients (15.2%). Ablation of the upper quartile of Ripple Frequency resulted in AF organization to a readily mappable rhythm (AT/AFL) in 28 patients (38.9%), and MAT in 25 patients (34.7%). Cardioversion was required in 28 patients (38.9%); 8 (11.1%) for AF, 16 (22.2%) for MAT, and 4 (5.6%) for refractory AFL. Table 2.

**Table 2.** Ablation Procedure Characteristics (n = 72)

| Variable | Mean ± SD / n (%) |
| --- | --- |
| Measured LA volume (mL) | 160.0 ± 39.4 |
| Baseline LA pressure (mmHg) | 13.4 ± 4.4 |
| AF pacing induction required | 18 (25.0) |
| Pulmonary vein isolation performed | 72 (100) |
| AF termination with PVI-only | 6 (8.3) |
| AF termination with ablation | 64 (88.9) |
| **Method of Termination** |  |

| **Variable** | **Mean ± SD / n (%)** |
| --- | --- |
| **PVI** | 6 (8.3) |
| **Ripple Frequency Ablation** | 58 (90.6) |
| **Cardioversion for AF only** | 8 (11.1) |
| **Total cardioversions performed** | 28 (38.9) |
| **Total fluoroscopy time (min)** | 0.48 ± 0.86 |
| **Total radiation dose (mGy)** | 0.35 ± 0.91 |
| **Total RF time (minutes)** | 35.2 ± 11.6 |
| **Total procedure time (min)** | 156.2 ± 35.0 |

The mean number of unique Ripple Frequency sites ablated per patient was 4.4 ± 2.4. The most frequently ablated non-PV sites are summarized in Table 3 and Figure 3.

**Figure 3:**
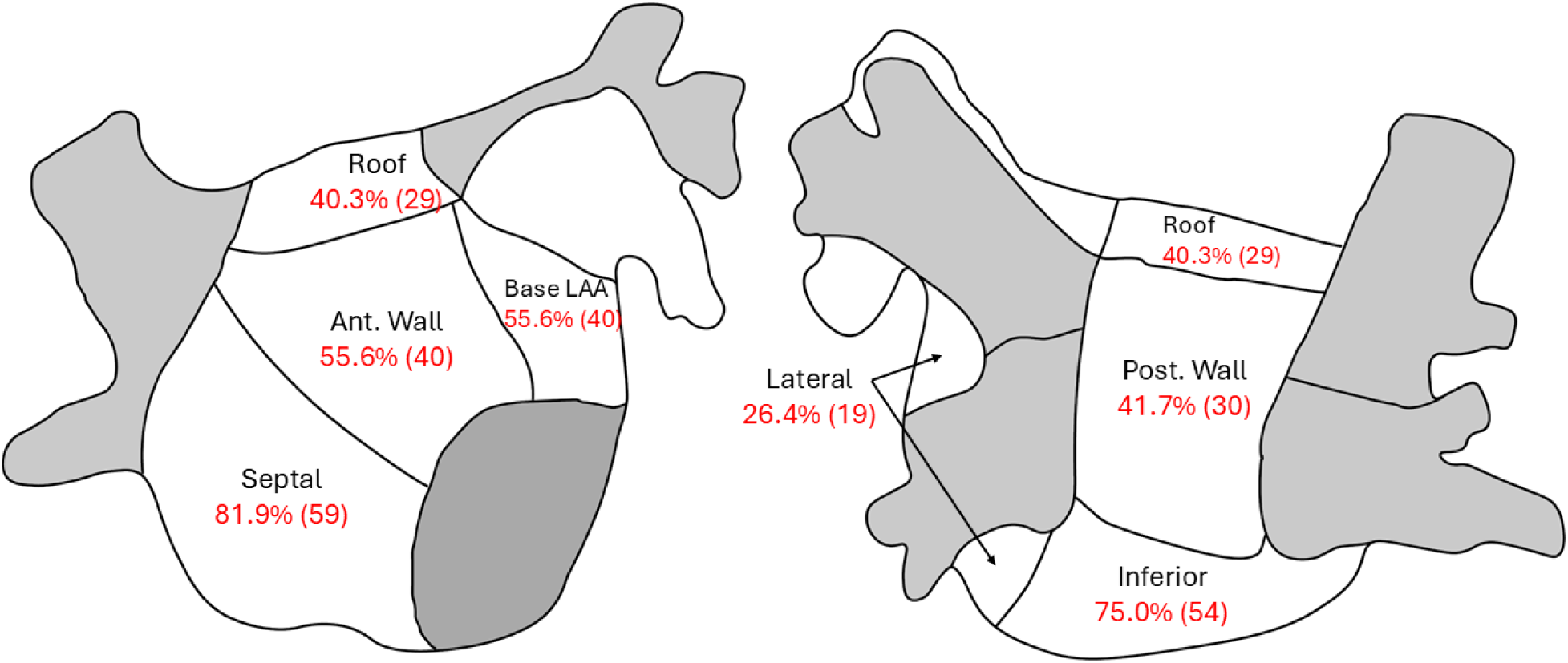
Left atrial ablated regions for Ripple Frequency. The left atrial shell is partitioned into eight anatomical segments—anterior wall, septum, roof, posterior wall, inferior wall, base of the left atrial appendage, peri-mitral isthmus, and left pulmonary-vein antrum. For each segment, the figure displays the percentage of procedures in which sites within the upper quartile of Ripple Frequency were identified and subsequently ablated.

**Table 3:** Ripple Frequency Guided Ablation Sites.

| **Atrial Region** | **n (%)** |
| --- | --- |
| **Septal LA** | 59 (81.9) |
| **Inferior LA** | 54 (75) |
| **Anterior LA** | 40 (55.6) |
| **Base of LAA** | 40 (55.6) |
| **Posterior LA** | 30 (41.7) |
| **Roof LA** | 29 (40.3) |
| **Lateral LA** | 19 (26.4) |
| **Coronary Sinus** | 18 (25) |
| **Right Atrial Appendage** | 12 (16.7) |
| **Posterior and Lateral RA** | 10 (13.9) |
| **Septal RA** | 4 (5.6) |

The mean total radiofrequency (RF) ablation time was 35.2 ± 11.6 minutes, with a mean procedure duration of 156.2 ± 35.0 minutes. (Table 2) No significant complications were observed (0 of 72 patients; 0.0 %, 95 % CI 0.0–4.1 %).

### 3.4 12-Month Arrhythmia Freedom Outcomes

After a 90-day blanking period, the Kaplan–Meier estimate of freedom from AF lasting >30 seconds at 12 months was 95.8% (69 of 72 patients; 95% CI 87.6-98.6). Freedom from AT/AFL was 76.4% (55 of 72; 95% CI 64.8–84.6), and freedom from any atrial arrhythmia was 72.2% (52 of 72; 95% CI 60.3-81.1). Thirteen patients (18.1%) underwent a second ablation procedure, resulting in a mean of 1.2 procedures per patient. Of these repeat procedures, 12 of 13 (92.3%) were for AT or AFL, and 1 of 13 (7.7%) for AF. Following the final procedure (i.e., one or two), freedom from AF increased to 97.2% (70 of 72; 95% CI 89.3-99.3), freedom from AT/AFL to 93.1% (67 of 72; 95% CI 84.1-97.0), and freedom from any atrial arrhythmia to 90.3% (65 of 72; 95% CI 80.7-95.2). At the 12-month follow-up, 24 of 72 patients (33.3%) were taking at least one class I or III AAD, most commonly sotalol (14 of 24) and flecainide (5 of 24). (Table 4, Figure 4)

**Figure 4:**
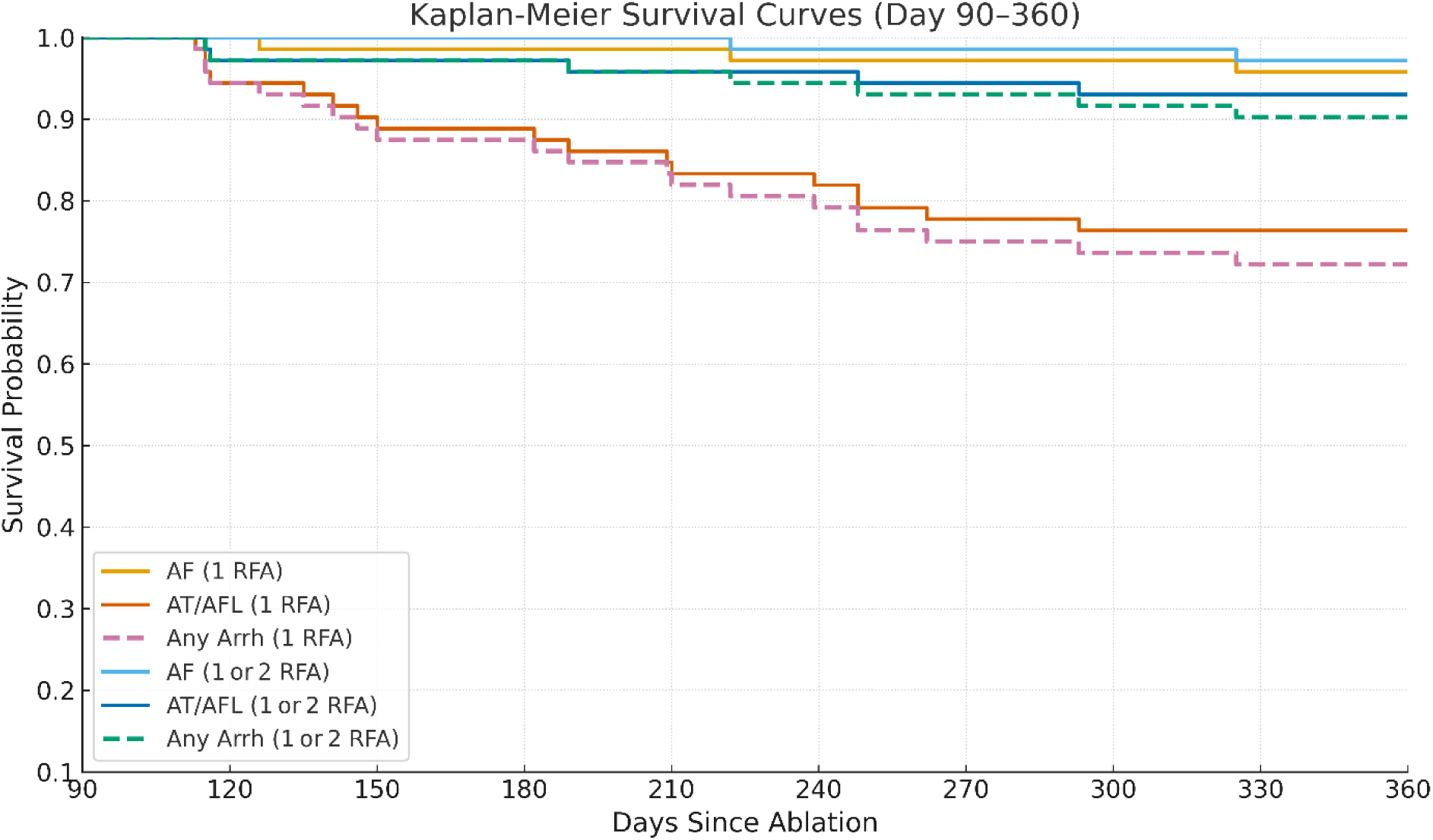
The Kaplan–Meier Survival Estimates for freedom from arrhythmia for AF, AT/AFL and any arrhythmia after a single procedure and after one or two procedures.

**Table 4.** Arrhythmia Freedom. After Single vs. Final Ablation. Values are Kaplan–Meier estimates at 12 months after a 90-day blanking period; 95 % confidence intervals calculated with Greenwood’s formula

| **Outcome** | **After Single Ablation n/N** | **% (95% CI)** | **After Final Ablation n/N** | **% (95 % CI)** |
| --- | --- | --- | --- | --- |
| **Freedom from AF recurrence** | 69 / 72 | 95.8%<br>(87.6-98.6) | 70 / 72 | 97.2%<br>(89.3-99.3) |
| **Freedom from AT/AFL** | 55 / 72 | 76.4%<br>(64.8-84.6) | 67 / 72 | 93.1%<br>(84.1-97.0) |
| **Freedom from any atrial arrhythmia** | 52 / 72 | 72.2%<br>(60.3-81.1) | 65 / 72 | 90.3%<br>(80.7-95.2) |

### 3.5 Comparison with benchmark PVI-only cohort

To provide clinical context, we compared our findings with the pulmonary-vein-isolation (PVI) arm of the STAR-AF II trial, in which 12-month AF freedom was 60 % (37/61). In our cohort, Kaplan–Meier estimated freedom from AF at 12 months was 95.8 % (69 of 72). The unadjusted absolute difference between the two cohorts was 35.8% (95% CI 22.3–40.0%). However, given differences in study era and rhythm monitoring intensity, this cross-study comparison is hypothesis-generating only and should not be interpreted as evidence of superiority.

## 4. Discussion

### Main Findings

This study evaluated the Ripple Frequency algorithm to identify non-PV AF sources for ablation in patients with persistent AF. The key findings were: 1) 95.8% of patients were free from AF recurrence at 12 months following a single procedure; 2) 90.3% were free from any atrial arrhythmia after a mean of 1.2 ablations; 3) acute AF termination was observed in 88.9% of patients; and 4) no major complications were observed. When benchmarked against the STAR-AF II PVI-only arm (60%), the unadjusted absolute difference in 12-month AF freedom was 35.8 percentage points; this cross-study comparison is hypothesis-generating only.

### Ripple Frequency Mapping

Ripple Frequency maps highlight atrial regions that exhibit sustained, spatially contiguous electrograms with repetitive high frequency dV/dt changes. Such frequent directional voltage changes may occur when multiple small wavefronts pivot, collide, or break up within the tissue. These patterns often reflect wavefront crowding, re-entry, or unstable conduction pathways that generate fragmented signals, features that have been associated with possible AF sources (9).

Since Ripple Frequency algorithms average regional clusters of neighboring electrodes and exclude points that fail voltage or point-density gates, the algorithm preferentially elevates regions where many electrodes register high-rate activity simultaneously. The most common sites observed with Ripple Frequency, including the inter-atrial septum and inferior left atrium, fulfill these criteria: both have relatively heterogeneous myocardium with abrupt anisotropic fiber transitions, dense autonomic innervation, and proximity to conduction bottlenecks (CS roof, fossa ovalis) that foster wavefront crowding, fragmentation, and stable high-frequency re-entry. (10, 11) Nevertheless, not all arrhythmogenic sources manifest these regional characteristics.

In contrast, focal AF sources (e.g., ligament of Marshall / mitral isthmus–CS–LA ridge complex) occupy only a few millimeters, fire intermittently, and may be epicardial or deeply intramural, so their electrograms are detected by at most one or two electrodes and are often suppressed by the algorithm’s amplitude and spatial-smoothing filters; consequently, they rarely rise into the upper quartile of Ripple Frequency rankings and may require complementary focal-trigger mapping techniques for reliable detection.

### Ripple Frequency Strategy

Ripple Frequency mapping was developed to identify AF sources. Unlike rotor ablation (12) and complex fractionated atrial electrogram (CFAE) ablation (2), Ripple Frequency highlights regions in which multiple electrograms within the predefined radius demonstrate high signal fragmentation in a 2.5 second acquisition window. By contrast, CFAE relies on features of a single electrogram (13), and rotor mapping emphasizes repetitive wavefront activation patterns (12). The first implementation of this technique used manual review of Carto 3 Ripple Maps to mark visual high-frequency ripple activation (HFRA) sites. As a proof of concept, in a comparative study, HFRA-guided ablation achieved higher AF termination (91.2% vs 52.4%; p<0.0001) and superior 18-month AF-free survival (98.2% vs 81.4%; p=0.005) compared with a standard stepwise approach (4,14).

To reduce subjectivity and streamline detection, the Ripple Frequency algorithm was developed. An initial evaluation using retrospective map analysis, the Ripple Frequency algorithm identified visually confirmed HFRA sites with 96.7% sensitivity and 91.1% specificity (5). AF terminated during ablation in 90.2% of these patients, and Ripple Frequency had an 86.5% specificity for colocation at the sites required for AF termination. One-year freedom from AF in this cohort was 92.9%, consistent with prior HFRA-guided outcomes (5). The results produced by the algorithm are verifiable by examining the electrogram markers directly; algorithm outputs include time- and location-synchronized electrogram markers that reference the raw signals. This permits independent verification and adjudication of detected AF sources, reducing reliance on opaque automated decisions.

Building on these Ripple Map and Ripple Frequency series, the current study evaluated real-time use of the Ripple Frequency map to guide ablation strategy. The findings support the hypothesis that atrial regions exhibiting high Ripple Frequency activity may mark clinically important sources in persistent AF. (4,5) There are several alternative mapping strategies which suggest a potential benefit for non-PV source AF mapping including Electrographic Flow Mapping which identified AF sources in 60% of patients, and when those sources were ablated, success improved to 95% (15). The UNCOVER AF trial demonstrated that noncontact charge density mapping was associated with significant improvements in arrhythmia-free survival (16), and ablation of spatiotemporal dispersion regions also improved AF freedom. (17) Finally, the TAILORED-AF trial found, after 12-months, 88% of patients in the tailored arm were free of AF compared with 70% of patients in the PVI-only arm (*P* < 0.0001). (18) Our work, combined with these studies, collectively supports a strategy of identifying and targeting non-PV AF sources to improve ablation outcomes in persistent AF.

### Comparison with Prior Experience

Support for non-PV sources remains conflicting, however, likely due to the variable methodologies utilized for identifying AF sources. Other ablation strategies using alternative AF source mapping, and particularly anatomic based ablation have yielded only modest or no benefit. For example, the STAR-AF II trial showed no significant improvement when linear lesions or CFAE ablations were added to PVI (2); rotor-based approaches did not demonstrate consistent advantages over PVI alone in REAFFIRM (11), and dominant-frequency mapping similarly demonstrated only modest improvement (18). In the CAPLA trial, posterior LA isolation was found to have no significant incremental benefit beyond PVI in persistent AF. (19) The SUPPRESS-AF trial likewise found that routine low-voltage area ablation following PVI failed to increase 12-month freedom from AF compared with PVI alone (61% vs 50%; p = 0.13). (20) These inconsistencies may reflect technical limitations in accurately identifying AF sources as well as the heterogenous nature of AF sources may not be well treated with anatomic, non-physiologic ablation strategies.

Ripple Frequency maps identified the posterior LA as an AF source in a minority of cases (41.7%), aligning with the CAPLA trial findings. Ripple Frequency sources were rarely observed in true low-voltage zones (< 0.1 mV) which is concordant with the SUPPRESS-AF findings. The Ripple Frequency algorithm applies multi-electrogram analysis to highlight regional fibrillatory electrograms in real time. The software provides real-time display with automated upper-quartile identification which yields patient-specific targets.

Prior work found the highest-quartile Ripple-frequency regions encompassed a relatively small portion of the total atrial surface area (5.6 ± 5.1%; range of 0.3% to 20.1%) (5); accordingly, the extent of ablation in this cohort was modest (mean RF time 35.2 ± 11.6 min) which may avoid unnecessary ablation, minimizing procedural risks and preserving atrial function.

In our experience, Ripple Frequency mapping combined well within existing workflows. No mapping- or ablation-related complications were observed, and procedure times and fluoroscopy exposure were consistent with standard persistent AF ablations, indicating minimal added complexity from this technique.

#### Limitations

This was a single-center observational study conducted by investigators involved in algorithm development, introducing potential bias. Automatic target generation and protocol adherence may have helped to mitigate—though not eliminate—this concern; multicenter validation is warranted. Prospective follow-up protocols were not standardized; rhythm-monitoring strategy was clinically determined and may have underestimated true recurrence. Outcomes were compared with an external historical cohort; differences in follow-up intensity, operator technique, and era may introduce residual confounding. Although Ripple Frequency mapping effectively identified and guided ablation of non-PV sources, analysis for AF-source mechanism was not performed. The cohort predominantly comprised patients with moderate left atrial enlargement (mean 46 mm) and preserved LVEF (mean 53.6%). These results therefore may not generalize to patients with severe atrial dilation or left ventricular dysfunction.

## 5. Conclusion

Patient-specific ablation guided by Ripple Frequency mapping was associated with high 12-month freedom from atrial fibrillation and no major complications. These findings support adjunctive ablation of residual AF sources after PVI. Multicenter trials are warranted to compare effectiveness with standard strategies. De-identified participant data and statistical code will be made available upon reasonable request to the corresponding author, following publication.

## Data Availability

De-identified participant data and statistical code will be made available upon reasonable request to the corresponding author, following publication.

## Abbreviation List

AF: atrial fibrillation
PVI: pulmonary vein isolation
PV: pulmonary vein
CFAE: complex fractionated atrial electrogram
HFRA: High-frequency Ripple activation
ECG: electrocardiogram
CIED: cardiac implantable electronic device
AT: atrial tachycardia
AFL: atrial flutter
LA: left atrium
RA: right atrium
MAT: multifocal atrial tachycardia
AAD: antiarrhythmic drug
CI: confidence interval
RD: risk difference
BMI: body mass index
LVEF: left ventricular ejection fraction
LAA: left atrial appendage
RF: radiofrequency
mGy: milligray

